# Kineret - The largest Israeli chain of hospitals’ data available now in OMOP common data model format enabling broad collaborative research

**DOI:** 10.1101/2025.07.04.25330663

**Authors:** Nadav Rappoport, Guy Livne, Naama Perry Cohen, Nir Makover, Hadas Eshel-Geva, Hadar Kapach, Tomer Hadad, Yarin Alon, Robyn Rubin, Segev Chai, Shirell Da Villa, Ohad Hochman

## Abstract

**Background:** In 2021, the Israeli Ministry of Health established the *Kineret* initiative for standardizing clinical data across its medical centers and making it accessible for secondary use, research, and development. The main objectives were to reduce burdens and bottlenecks in data extraction, data cleaning, and data sharing in order to enable the reuse of patient data for translational research. The Directorate of Government Medical Centers is the governing body for the network of government healthcare centers, which includes 25 medical centers spread across Israel with 11 general medical centers, 9 mental health centers, and 5 geriatric healthcare centers.

**Methods:** After considering several alternatives, the Observational Medical Outcomes Partnership Common Data Model (OMOP CDM) was chosen as the data standard for harmonizing the terminologies across institutions. An instance of ATLAS, an open-source software for scientific analyses, was made available for researchers on a secure cloud for researchers within the medical centers. This enables self-feasibility tests and exploratory analysis in an efficient way. Approved studies are conducted in a secure virtual environment on a cloud-based platform, which provides flexibility for a variety of computational resource needs.

**Results:** Since the initiation of the project, six medical centers have joined *Kineret* and their data has been fully integrated into the data lake. The seventh medical center is expected to be fully integrated during 2025.

**Conclusion:** *Kineret* enables not only internal multi-site studies but also international studies with ease. We see that *Kineret* enables studies for improving local and worldwide health and healthcare. Data description is available at https://kineret.health.gov.il/

## Introduction

In recent years, the medical research landscape has undergone a significant transformation, with data sharing and collaborative learning becoming central to advancing healthcare knowledge and outcomes. The integration of hospital data into research databases is a critical step in this process, enabling researchers to harness vast amounts of clinical information for analysis and discovery [1]. However, the heterogeneity of healthcare data systems across institutions creates significant challenges for data harmonization, collaboration, and comparative studies [2]. Moreover, medical centers face substantial operational bottlenecks that delay studies. Limited technical resources and insufficient specialized personnel trained in clinical data extraction frequently resulted in extended delays, with investigators often waiting a long time to access relevant clinical information from the electronic health record systems or even abandoning studies. These constraints particularly affected cross-institutional research, though even single-site studies suffered from inefficiencies as investigators depended on overburdened informatics teams for relatively straightforward data queries [3].

To address this challenge, adopting international common data models has gained traction in the medical research community. These standardized frameworks provide a uniform structure for organizing and representing healthcare data, facilitating interoperability and simplifying collaboration across institutions worldwide. Several common clinical data models have been developed and implemented, including the Observational Medical Outcomes Partnership Common Data Model (OMOP CDM) [4], the Patient-Centered Outcomes Research Network (PCORnet) Common Data Model, the Sentinel Common Data Model [5], and the i2b2 (Informatics for Integrating Biology and the Bedside) data model [6]. Each CDM has advantages and limitations [7–10].

Among these, the OMOP CDM, developed by the Observational Health Data Sciences and Informatics (OHDSI) collaborative, has gained widespread adoption. OHDSI, an international network of researchers and observational health databases, aims to improve health by empowering a community to collaboratively generate the evidence that promotes better health decisions and better care. The OMOP CDM standardizes the format and content of observational data, enabling large-scale analytics and facilitating the development and application of reliable evidence-based algorithms.

Governmental healthcare data represent a vast and valuable resource for medical research, public health initiatives, and health policy development. However, these data are often stored in disparate systems with varying structures and terminologies, making it challenging to conduct comprehensive analyses or compare findings across different regions or countries. The process of converting governmental healthcare data into a standardized format, such as the OMOP CDM, holds immense significance. This transformation unlocks the potential for more robust and extensive studies and fosters international collaborations.

There is the advantage of transforming institute-wide data to OMOP CDM and not only on a per-study/per-cohort basis. As once the data is transformed once, it can be used for multiple studies, in multiple clinical fields [11–13]. Several country-wide efforts have been reported to transform and standardize EHRs to the OMOP CDM. In Estonia, a national effort was made to transform EHR data from three national health databases [14]. In the UK, a large-scale transformation of linked EHR data from multiple national sources to OMOP CDM was undertaken to convert over 1 billion rows of data from over 216 million encounters across three EHR sources [15].

In Israel, the Ministry of Health (MOH) regulates health services. In addition, MOH owns and managed the largest network of hospitals in Israel through the Directorate of Government Medical Centers established in 2015. The networks composed of 11 generic hospitals (Barzilai, Bnei-Zion, Hille-Yafe, Wolfson, Ziv, Galil, Tzafon, Rambam, Sheba, Shamir, Ichilov), 8 Psychiatric centers (Abarbanel, Beer-Yaakov, Beer-Sheva, Jerusalem Center, Mazor, Karmel, Lev-Sharon, Shaar-Menashe), and 5 Geriatric (Dorot, Fleeman, Rishon-Lezion, Shoham, Shmuel Harofe). In 2019, the Directorate of Government Medical Centers launched an initiative to empower research and innovation using data from its hospitals. The name of the initiative is *Kineret*, which, on the one hand, is the Hebrew name for the sea of Galilee, representing the data lake. On the other hand, *Kineret* is also an abbreviation in Hebrew for medical data mining for insights 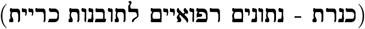 For this purpose, the Directorate of Government Medical Centers developed a pipeline for the ETL (Extract Transform Load) process in collaboration with the hospitals.

The extraction, transformation, and loading (ETL) process presented several unique challenges that distinguished this implementation from typical data harmonization efforts. Unlike projects focused on specific cohorts or clinical departments, this initiative encompassed comprehensive electronic health records (EHR) representing the full spectrum of clinical services provided by Israeli hospitals, including emergency care, inpatient treatment, and ambulatory services. The heterogeneity of source data systems—including Chameleon, NAMER, Picture Archiving and Communication System (PACS), and additional platforms—significantly increased the technical complexity of the integration process. Further complicating the ETL workflow was the prevalence of Hebrew-language clinical documentation without complete mapping to international standard vocabularies, necessitating extensive cross-language terminology harmonization. To optimize computational resource allocation, the implementation strategy employed a hybrid processing architecture wherein only essential operations were performed on-premise, with the majority of data processing occurring in cloud-based environments. This architectural decision subsequently necessitated adaptation of existing regulatory frameworks to accommodate secure cloud-based storage and processing of protected health information within the constraints of Israeli healthcare governance standards.

This manuscript describes the methodological approach and technical considerations involved in transferring governmental healthcare data into the OMOP CDM v5.3. By elucidating this process, we aim to provide a blueprint for other institutions and governmental bodies seeking to enhance the utility and accessibility of their healthcare data for research purposes. Furthermore, this work contributes to the broader goal of creating a global network of standardized healthcare databases, ultimately accelerating the pace of medical discovery and improving patient care.

## Methods

The design of the ETL process and its implementation was composed of several steps. First, data profiling was executed. Then, characterization and definition of the transformation process was made by experts in the clinical notes and coders. Based on their documentation and specifications, a team of data engineers and dev-ops implemented the transformation and the load process. Tests were implemented along the process. Tests were composed of logical and quantitative tests to ensure that no records were missed or duplicated. The standard tests we used are the Data Quality Dashboard (DQD)^1^ and Achilles^2^ from OHDSI. On top of that, we add many more tests. For example, a quantity comparison of the source data and the target data.

### Care Sites’ Legacy EHR Systems

Each medical center in the network has its own instance of EHR system. They were all historically based on NAMER as the main EHR system, and in the last 4 years, gradually switched to Chameleon. Other EHR systems include MAX, Clicks, and Labs. Although the base infrastructures and EHR systems are managed on the network-level, yet local adaptation and customization are made. Therefore, the extraction part was mostly shared, but we had to make adaptations for site-specific systems.

### Extract, Transform, Load Process

The ETL process for transferring clinical data from a hospital’s on-premise legacy system to the OMOP CDM comprised several pivotal stages to guarantee both compliance with data protection regulations and the integrity of the data. Initially, the data extraction was conducted on-premise within each hospital, where sensitive patient information was de-identified in accordance with stringent local privacy standards. This step ensured that no identifiable information was exposed during subsequent processing. Each individual receives a hashed, de-identified id that is consistent across all tables and databases. This consistency is possible as patients in Israel are identified by their governmental identification number (similar to the United States’ social security number). The newly introduced unique patient identifier replaces the actual identifier in each table consistently. Other identifier values, such as phone numbers and addresses, are completely removed on-premise before data is uploaded to the cloud.

Once de-identified, the data underwent a series of transformations to align with the OMOP CDM’s schema, including mapping local coding systems to standardized terminologies, restructuring data tables, and ensuring consistency and completeness across datasets. These transformations were performed in a secure cloud environment, which provided the necessary computational resources and security measures to handle large-scale clinical data. Finally, the transformed data was loaded into a data base in OMOP CDM format, enabling seamless integration with other datasets and facilitating advanced analytics and research within the secure cloud infrastructure.

### Unique ID solution

A significant challenge in multi-institutional healthcare networks, such as the Israeli Ministry of Health system, is the secure linkage of patient records across different care providers. To address this challenge, we implemented a Generated Key (GK) methodology for patient identification. The GK approach enables the creation of unique patient identifiers by combining multiple data sources from disparate legacy systems into unified OMOP CDM tables. This mechanism was successfully employed to generate all entity identifiers within the CDM, ultimately enabling the integration of data from six hospitals into a single OMOP CDM instance, thereby facilitating multi-center research studies. The GK implementation ensures patient privacy while maintaining the ability to conduct comprehensive cross-institutional analyses.

### Periodical Updates

The maintenance of the OMOP CDM requires a comprehensive refresh strategy that differs from traditional data warehouse approaches. We implemented periodic full-load updates of the entire database for two critical reasons. First, the OHDSI community regularly updates the ATHENA vocabulary mappings, necessitating complete data remapping to maintain standardization. Second, source medical records frequently undergo retrospective modifications, with changes occurring days or even months after initial documentation. These retrospective modifications can have cascading effects beyond the immediately affected records. To address these challenges, we developed an automated ETL pipeline that performs complete data refresh cycles on a periodic basis, including comprehensive quality assurance testing. This approach ensures that any modifications to the ETL process, such as the mapping of newly encountered source codes, are uniformly applied across the entire historical dataset rather than being limited to recent records. This methodology maintains data consistency and standardization across the temporal spectrum of our clinical data.

### Terminologies mapping

The standardization of medical terminologies across multiple healthcare centers presented several significant challenges in our OMOP CDM implementation: First, the Israeli Ministry of Health has established a local catalog called IC, which extends the ICD9-CM diagnosis coding system. This proprietary extension is not included in the ATHENA standardized vocabularies and cannot be processed through USAGI, the OHDSI terminology mapping tool [16]. Second, individual hospitals maintain the authority to modify and expand their local vocabularies. Third, a portion of the codes and their descriptions are documented in Hebrew, adding a layer of complexity to the standardization process. To address these challenges, we implemented a two-phase mapping approach. In the initial phase, we developed a language-based model to establish correspondences between each medical center’s vocabulary codes and descriptions with OMOP CDM standard concepts. This automated approach achieved a mapping accuracy exceeding 0.9 for approximately 85% of the vocabulary terms. For the remaining 15% of terms, we employed a manual mapping process utilizing domain experts and the USAGI tool, developed by the OHDSI collaborative [16]. USAGI facilitates the conversion of data warehouse terminology to standardized concepts, but requires expert oversight to ensure accurate mapping. We strategically engaged domain specialists: laboratory professionals for laboratory terms, pharmacists for medication-related terminology, and physicians for diagnostic codes. Each specialist independently conducted mappings within their respective domains of expertise.

Through this combined approach, we successfully mapped over 64,000 source concepts to approximately 33,000 standard target concepts (Table 1). The condition and procedure catalogs comprised the largest proportion of source concepts. While the medical centers largely shared common source codes, each facility maintained unique extensions to address local requirements (Table 2).

**Table 1.** Summary of source vocabularies and number of unique source codes in each and the number of unique mapped target codes.

|  | Source vocabulary ID | #codes in source | #mapped target codes |
| --- | --- | --- | --- |
| 1 | ANTIBIOTIC_SENSITIVITY | 4 | 4 |
| 2 | AN_MOH | 343 | 77 |
| 3 | CHAMELEON_ALLERGIES | 56 | 27 |
| 4 | CHAMELEON_CONDITION_STATUS | 10 | 2 |
| 5 | CHAMELEON_DRUGS | 3661 | 2766 |
| 6 | CHAMELEON_DRUG_SENSITIVITY | 328 | 141 |
| 7 | CHAMELEON_HABIES | 5 | 5 |
| 8 | CHAMELEON_MED_ROUTE | 87 | 36 |
| 9 | CHAMELEON_SCORE | 51 | 45 |
| 10 | CHAMELEON_SURG_ROLE | 7 | 5 |
| 11 | CHAMELEON_TIME_STAMP | 3 | 3 |
| 12 | CHAMELEON_TPN | 615 | 632 |
| 13 | CHAMELEON_VITAL_SIGNS | 315 | 265 |
| 14 | COUNTRY_OF_ORIGIN | 237 | 227 |
| 15 | MARTIAL_STATUS | 5 | 5 |
| 16 | MAX_VITAL_SIGNS | 4 | 4 |
| 17 | MICRO_CULTURE_RESULTS | 2 | 14 |
| 18 | NAMER | 9 | 9 |
| 19 | NAMER_ADMITTED_FROM | 61 | 19 |
| 20 | NAMER_ANTIBIOTIC_SUSCEPTIBILITY | 151 | 137 |
| 21 | NAMER_CATALOG_1 | 10666 | 1392 |
| 22 | NAMER_CONDITION | 17207 | 10361 |
| 23 | NAMER_CONDITION_STATUS | 5 | 5 |
| 24 | NAMER_DEPARTMENTS | 1852 | 164 |
| 25 | NAMER_DISCHARGE_TO | 17 | 5 |
| 26 | NAMER_DISCHARGE_TO_MOVEMENT | 6 | 1 |
| 27 | NAMER_DISCHARGE_TO_RELEASE | 16 | 10 |
| 28 | NAMER_DRUGS | 6734 | 4410 |
| 29 | NAMER_GENDER | 2 | 2 |
| 30 | NAMER_HY | 52 | 44 |
| 31 | NAMER_LABS | 6633 | 3307 |
| 32 | NAMER_LABS_VALUE | 2 | 2 |
| 33 | NAMER_LAB_SPECIMEN | 360 | 60 |
| 34 | NAMER_LOCATIONS | 1139 | 261 |
| 35 | NAMER_MED_ROUTE | 81 | 45 |
| 36 | NAMER_MODIFIERS | 14 | 3 |
| 37 | NAMER_OPERATOR | 5 | 5 |
| 38 | NAMER_PROCEDURES | 11725 | 6746 |
| 39 | NAMER_RISK_FACTORS | 363 | 326 |
| 40 | NAMER_TASKS | 208 | 41 |
| 41 | NAMER_TIMESTAMPS | 29 | 16 |
| 42 | NAMER_TPN | 321 | 277 |
| 43 | NAMER_VISIT_DETAIL_ADMITTING | 15 | 3 |
| 44 | NAMER_VISIT_DETAIL_DISCHARGE | 8 | 6 |
| 45 | NAMER_VISIT_DETAIL_SOURCE | 7 | 5 |
| 46 | NAMER_VITAL_SIGNS | 63 | 36 |
| 47 | NAMER_ZBACTERIUM | 683 | 631 |
| 48 | NURSE_SPECIALITY | 23 | 13 |
| 49 | OTHER_SPECIALITIES | 251 | 75 |
| 50 | PHYSICIAN_SPECIALITIES | 60 | 57 |
| 51 | RELIGION | 7 | 5 |
| 52 | ROLE_MOH | 23 | 14 |
| 53 | UNIT_CONCEPT | 232 | 109 |

**Table 2.** Summary of the number of unique source codes and mapped target codes by medical center.

| Medical center | #unique source codes | #unique target codes | #unmapped source codes |
| --- | --- | --- | --- |
| Barzilai | 54766 | 27949 | 14 |
| Bnai-Zion | 55222 | 28202 | 13 |
| Hillel Yaffe | 55680 | 28037 | 13 |
| Nahariya | 54901 | 27994 | 13 |
| Tzfon | 56398 | 28318 | 13 |
| Shamir | 56097 | 28351 | 13 |

#### Diagnoses

The source vocabulary is based on ICD-9-CM, in addition to local modifications and additions. Therefore, the source ICD-9-CM concepts were mapped using the mapping tables available in Athena [17]. The rest of the terms were mapped by experts using USAGI [16].

#### Procedures

The source vocabularies of the procedures are CPT-4 and ICD-9CM. Therefore, the mapping from source concepts to target standards concepts was performed using the mapping tables from Athena [17].

#### Labs

The source vocabulary of the laboratory test names is LOINC and, therefore, was mapped using the mapping table from Athena [17]. The modifiers and units concepts were manually mapped to standard concepts using Athena.

#### Medications

The source vocabulary of medications is a local catalog. Moreover, many drugs have a commercial Israeli brand name which is not part of any international vocabulary.

Therefore, most drugs’ mapping was conducted using our automated approach. The non-mapped drugs were manually mapped to the standard vocabulary RxNorm [18] by pharmacists according to active ingredients, ATC concepts, and routes of administration. Routes of administration were manually mapped to standard concepts using USAGI to fill in the *route concept id* field.

#### Other Concepts

Other concepts that are originally in Hebrew like modifiers (like bilateral procedure) and types (like *visit type concept id* and *condition type concept id* ) were manually mapped by querying Athena.

### Non-OMOP data

One limitation of OMOP CDM is that it is a structured database which does not cover all data available in the EHR. For example, images, ultrasound recordings, PDF files, ECG, and fetal monitor signals. In order to enrich *Kineret* datalake we created processes that extract such types of data from the source systems. The non-OMOP items are automatically assigned to the appropriate de-identified visits and patients, so the data is enriched appropriately. This data is not periodically updated, but rather extracted upon request and approval of studies.

### Provision of a Virtual Research Machine

The OMOP data of each medical center is uploaded to an individual PostgreSQL instance on Amazon Web Services. One ATLAS [19] instance on the cloud is connected to the individual databases (Fig 1).

**Fig 1.**
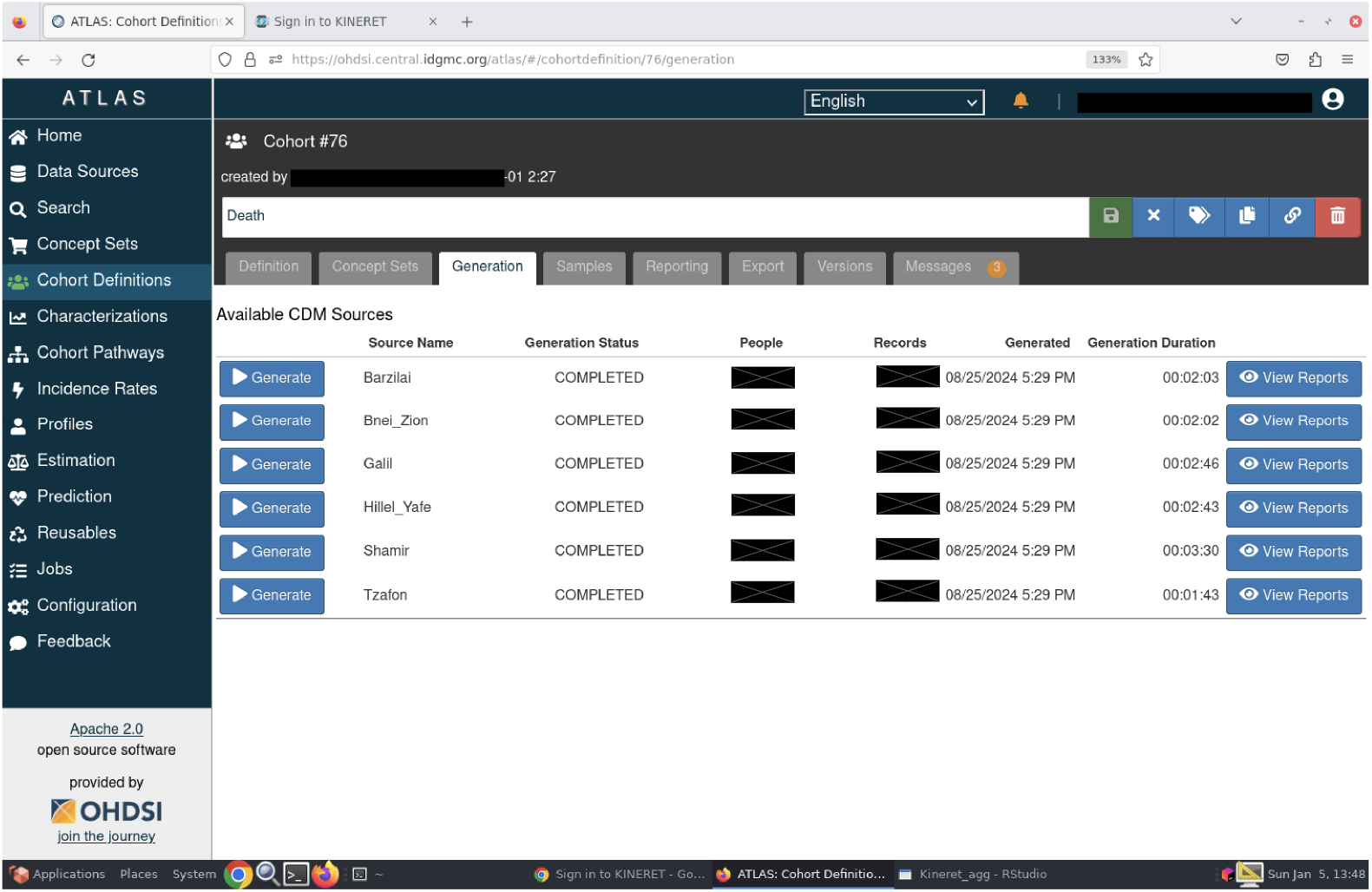
Screenshot of the centralized ATLAS instance of *Kineret*. A screenshot from the Cohort Definition tab showing the different available hospitals.

Once a data application is approved by the IRB committee and the PIs had signed the agreement with *Kineret*, they get a virtual machine for analysis and development with the approved data.

## Results

ETL process was developed and executed in multiple rounds for enhancing the correctness and completeness of the process as well as improving the quality of the data. A summary of the row numbers in the OMOP database is given in Table 3.

**Table 3.** Number of records per OMOP CDM table.

| Table | #Rows |
| --- | --- |
| care_site | 1,263 |
| conditions | 70,123,985 |
| death | 603,271 |
| device_exposure | 3,613,875 |
| drug_exposure | 97,626,180 |
| measurements | 1,358,142,289 |
| observation | 38,794,747 |
| persons | 6,836,687 |
| procedures | 82,888,358 |
| provider | 47,541 |
| specimen | 51,436,729 |
| visits | 52,289,122 |
| visit_details | 109,964,822 |

In total, in the mapped data there were 3,287,140 female patients, 3,539,384 male patients, and 10,163 others (Table 4). The distribution of year of birth of patients in *Kineret* is provided in Fig 2.

**Table 4.** Number or persons per sex.

| Sex | #Persons |
| --- | --- |
| FEMALE | 3,287,140 |
| MALE | 3,539,384 |
| No matching concept | 10,163 |

**Fig 2.**
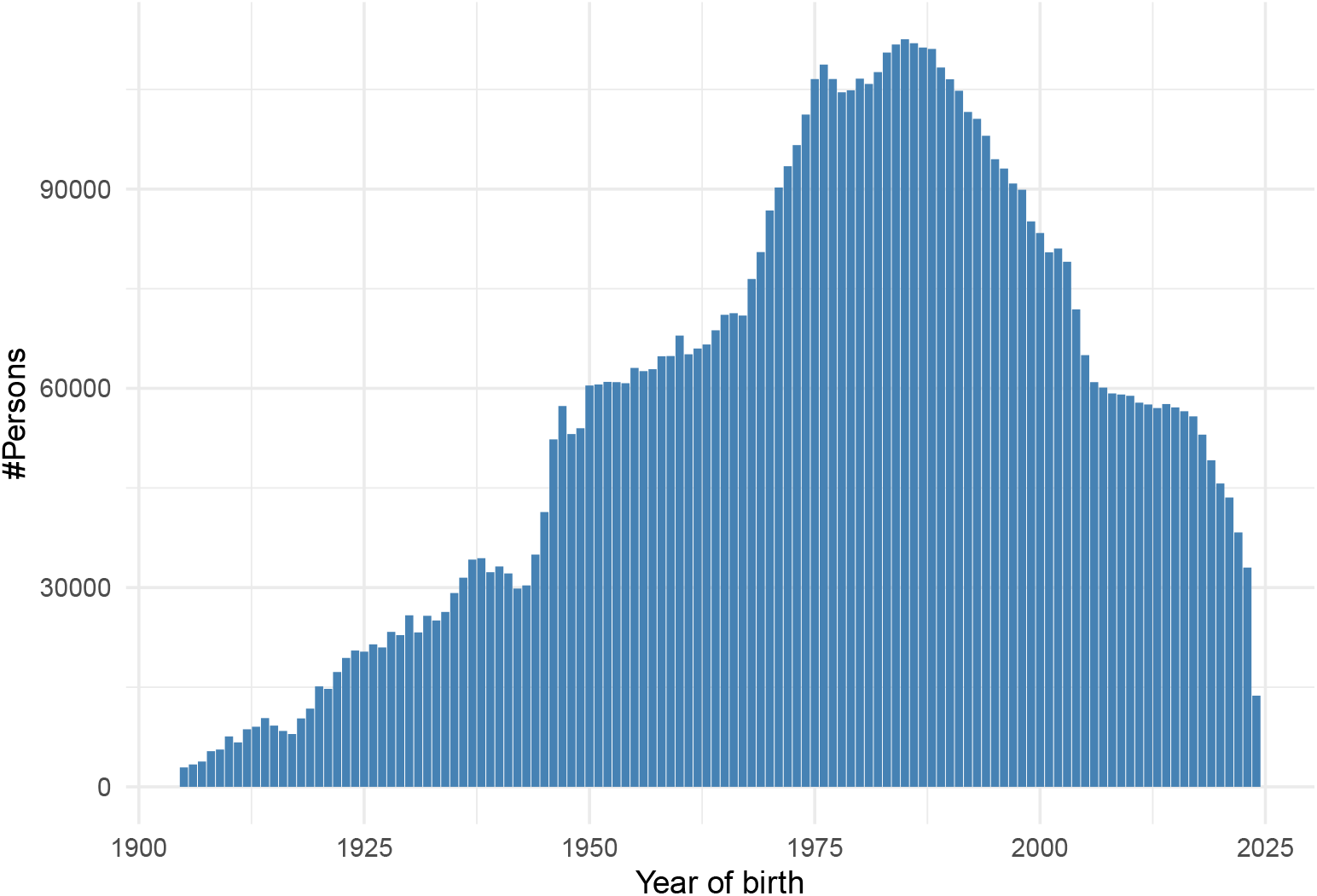
Number of persons by year of birth. Histogram of number of persons born in every year. from 1900 to 2025.

### Visit types

The medical centers in the network provide inpatient and outpatient care. Most visits are outpatient visits (about 29M, 55%), then emergency room visits (15M visits, 28%), and the rest are inpatient visits (8M, 16%) (Table 5).

**Table 5.** Number of records per visit type.

| Visit type | #Rows |
| --- | --- |
| Emergency Room Visit | 14,786,492 |
| Inpatient Visit | 8,537,702 |
| No matching concept | 2 |
| Outpatient Visit | 28,964,926 |

### Death records

Death records are updated in the source from the governmental Population and Immigration Authority. In this way, death records are updated even when a patient passes away not during a visit. Most death records (78%) in our datalake are from governmental reports (Table 6).

**Table 6.** Number of records per death type.

| Death record type | #Persons |
| --- | --- |
| EHR discharge summary | 84,241 |
| Government report | 472,274 |
| No matching concept | 46,756 |

EHR - Electronic Health Records.

## Discussion

The *Kineret* initiative represents a significant advancement in healthcare data standardization and research facilitation within Israel’s medical ecosystem. By establishing a centralized data platform in the OMOP Common Data Model format, *Kineret* addresses several fundamental challenges in clinical research while creating new opportunities for collaboration and discovery.

### Research Access and Workflow

The process for researchers to access *Kineret* data has been streamlined to balance efficiency with appropriate oversight (Fig 3). Internal researchers can begin with feasibility testing through the ATLAS interface, allowing them to refine research questions and determine appropriate cohorts before formal study initiation. External researcher, need to contact *Kineret* to perform the feasibility test. They can provide a detailed description of the cohort, or build on their own using a public ATALS instance and share the export JSON file. This preliminary step reduces the resources expended on studies that might otherwise prove infeasible due to insufficient sample sizes or data availability. Once a researcher identifies a viable study, they submit an application to the relevant Institutional Review Board (IRB). A notable regulatory advancement has transformed this previously cumbersome process. Previously, multi-center studies required separate IRB applications at each participating institution, resulting in redundant fees, administrative burden, and significant delays. The new streamlined approach allows researchers to submit a single primary IRB application, with other centers able to ratify this approval through an expedited internal process. This regulatory innovation significantly reduces barriers to multi-center research and promotes more collaborative investigations. Upon IRB approval and execution of the *Kineret* data use agreement, researchers receive access to a secure virtual machine with the approved dataset. This environment provides the necessary analytical tools while maintaining appropriate data governance and security controls. The cloud-based infrastructure offers scalability to accommodate varying computational requirements across different research projects, from small observational studies to complex machine learning applications.

**Fig 3.**
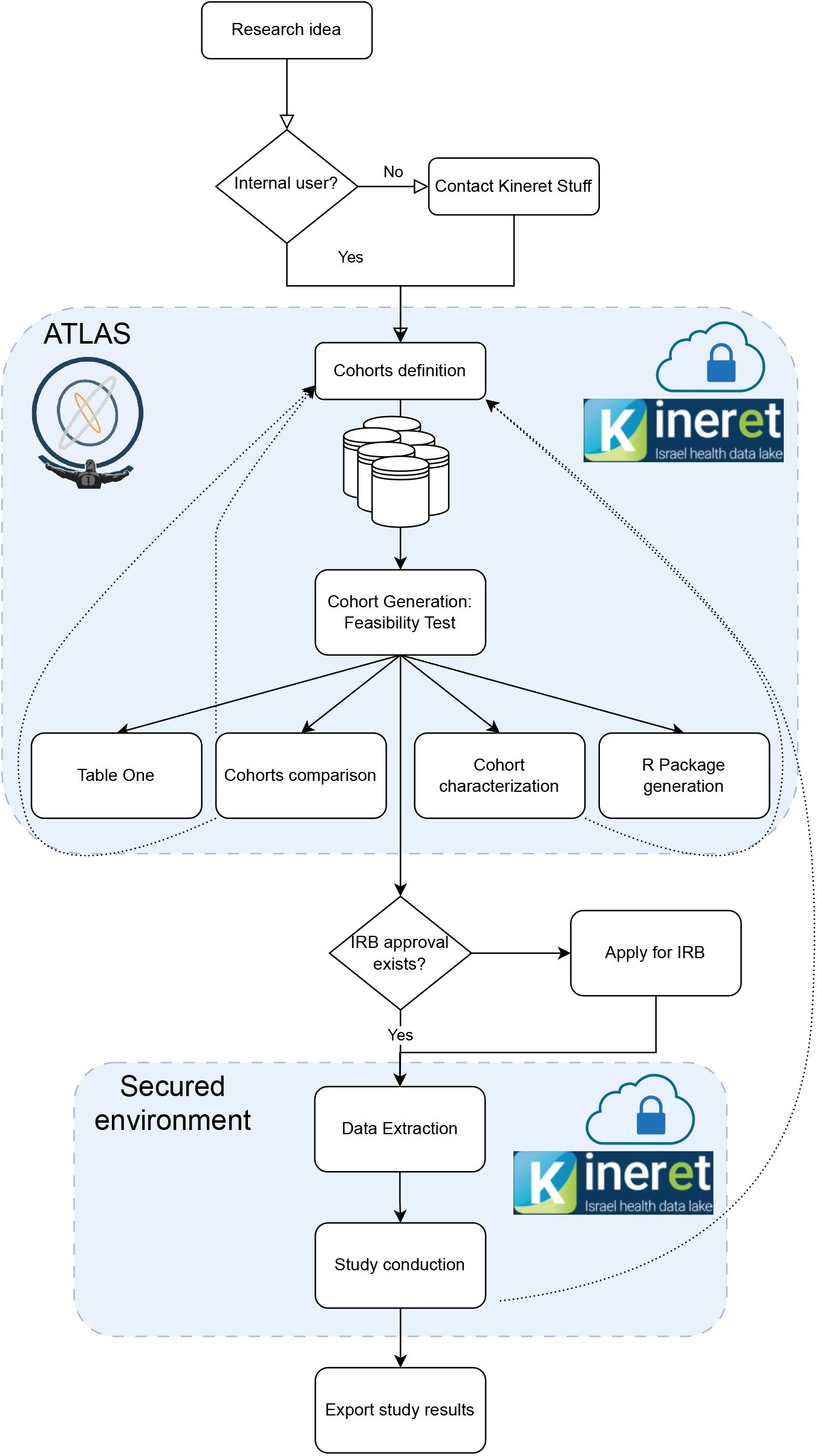
Illustration of process flow in *Kineret*. Internal users can access a secured environment with an ATLAS instance to generate cohorts from all sources at once or independently. To extract a cohort’s data and upload it to the secured study room, IRB approval is required. *Kineret* stuff supports every step if required by the user.

### Technical Infrastructure and Tools Integration

*Kineret* ‘s implementation has benefited substantially from integration with open-source tools developed by the OHDSI community and other stakeholders. These tools support the entire research lifecycle. USAGI facilitates concept mapping between local terminologies and standard vocabularies, addressing the particular challenges of Hebrew-language clinical documentation and proprietary extensions to standard coding systems. DQD and Achilles provide comprehensive quality assurance, enabling continuous monitoring and improvement of data quality metrics. ATLAS and PatientExploreR [20] offer intuitive interfaces for cohort definition, characterization, and exploratory analysis, allowing researchers to interact with the data without requiring advanced programming skills.

The platform’s cloud-based architecture offers several advantages beyond mere technical convenience. It enables secure access from any location, facilitating collaboration between researchers at different institutions and even international partnerships. The scalable infrastructure accommodates the growing volume of clinical data and increasing computational demands for advanced analytics, without requiring substantial capital investments in local hardware.

### Impact on Research and Clinical Practice

Since its inception, *Kineret* has supported many local and international studies that have yielded significant findings [21–23]. The standardized data structure has proven particularly valuable for cross-institutional comparisons, enabling investigations into practice variations and outcomes across different healthcare settings within the Israeli healthcare system. The initiative also addresses a critical bottleneck in clinical research: the time and technical expertise required to extract, clean, and prepare data for analysis. By providing pre-processed, standardized data, *Kineret* allows clinical researchers to focus on hypothesis generation and testing rather than data engineering. This shift has the potential to accelerate the translation of clinical insights into practice improvements. Furthermore, the OMOP CDM implementation facilitates participation in international research networks and consortia, positioning Israeli healthcare institutions within the global research community. This integration enables Israeli researchers to contribute to and benefit from large-scale, multinational observational studies that would be infeasible within a single healthcare system.

### Collaborative Work

As noted, the *Kineret* initiative was built upon the formation of a broad coalition comprising eight different entities — seven independent medical centers and the directorate of government medical centers that oversees them. Establishing this partnership required numerous meetings and discussions with hospital CEOs, financial officers, medical directors, and administrative managers. Each institution had its own concerns and resistance, necessitating a high degree of creativity to navigate the complexities.

As part of the solution, it was decided to establish a governance structure that included representatives from each hospital. Two dedicated forums were created: one focused on financial regulation, known as the Economic Regulation Committee, and another addressing clinical aspects, called the Research Forum. Each forum was staffed with appropriate representatives whose primary roles were to make joint decisions and advance the initiative both collectively and at an institutional level. This structured approach was crucial in fostering trust among all stakeholders.

At the outset, strict boundaries were set between the data lakes of each organization, ensuring full segregation—each internal researcher could access only their own hospital’s data. However, after approximately a year and a half of collaboration and trust-building, all institutions approved expanding access, allowing internal researchers at one medical center to view data from across the entire network.

### Challenges and Limitations

Despite its successes, the *Kineret* initiative has encountered several challenges worth noting. The standardization of terminologies across different healthcare settings required significant investment in automated mapping tools and expert review. While our approach achieved high mapping accuracy for most terms, approximately 15% required manual curation, highlighting the complexity of terminology harmonization across linguistic and institutional boundaries. One limitation of the OMOP CDM is its focus on structured data, which may not fully capture the richness of clinical information contained in unstructured notes, images, and other multimedia data. To address this limitation, we developed complementary processes to extract and link non-OMOP data types with patient records, providing a more comprehensive view of clinical information. However, these unstructured data elements present ongoing challenges for de-identification, standardization and analysis. The quarterly full-load update strategy, while ensuring data consistency, introduces computational overhead and temporary unavailability during update cycles. As the volume of data continues to grow, optimizing this process while maintaining data integrity will remain an important consideration.

### Future Directions

The *Kineret* initiative continues to evolve along several trajectories. Expansion to include additional medical centers within the Israeli healthcare system remains a primary goal, with the seventh medical center expected to be fully integrated during 2025. This expansion will further enhance the representativeness and statistical power of the dataset. Future development will focus on integrating advanced analytical capabilities directly within the *Kineret* platform, including natural language processing tools for unstructured clinical notes and machine learning frameworks for predictive modeling. These enhancements will enable researchers to leverage both structured and unstructured data for comprehensive clinical investigations. We also anticipate deeper integration with international research networks through the OHDSI consortium and the European Health Data & Evidence Network (EHDEN). These collaborations will facilitate participation in large-scale, distributed research studies addressing global health challenges.

## Conclusion

In conclusion, the *Kineret* initiative represents a significant advancement in the standardization and accessibility of healthcare data within Israel’s medical landscape. By adopting the OMOP CDM, *Kineret* has effectively transformed disparate clinical data from multiple medical centers into a unified format, facilitating enhanced research capabilities and collaborative learning. This initiative not only streamlines data extraction and analysis but also supports both national and international multi-site studies, thereby broadening the scope of potential research endeavors. The successful integration of data from various healthcare institutions underscores the importance of standardized frameworks in overcoming challenges related to data heterogeneity and interoperability. As *Kineret* continues to evolve, it promises to serve as a vital resource for advancing medical research, improving patient care, and fostering innovation within the healthcare sector. Furthermore, the experiences and methodologies developed through *Kineret* provide a valuable blueprint for other institutions and countries aiming to enhance their healthcare data infrastructures, thereby contributing to a global network of standardized healthcare databases that can accelerate medical discovery and improve health outcomes.

## Data availability

Aggregated data is available in the *Kineret* portal https://kineret.health.gov.il. Raw data is available upon IRB approval and a signed agreement.

## Acknowledgments

We would like to acknowledge all *Kineret* team at the Directorate of Government Medical Centers, Israeli Ministry of Health. Specifically, Dr. Orly Weinstein’s contribution in the first steps of this initiative. The steering committee of *Kineret* : Prof. Amos Katz, Prof. Mati Berkovitch, Prof. Zehava Vadasz, Prof. Avi Peretz, Dr. Khetam Hussein, Dr. Dikla Dahan Shriki, and Prof. Aviram Nissan. We would like to thank the chairs of the institutional IRB committees: Prof. Amos Katz, Prof. Mati Berkovitch, Prof. Kamal Hussein, Dr. Noa Brar-Yannai, Dr. Efrat Wolfowitz, and Dr. Einav Yefet.

## Footnotes

1 https://github.com/OHDSI/DQD

2 https://github.com/OHDSI/Achilles

## Notes

### Competing Interest Statement

The authors have declared no competing interest.

### Funding Statement

This study was funded by the Israeli authority of Innovation.

### Author Declarations

This manuscript does not report on a study involving human subjects, human data analysis, or clinical outcomes research. It describes the technical and methodological process of transforming structured clinical data from multiple Israeli governmental hospitals into the Observational Medical Outcomes Partnership (OMOP) Common Data Model (CDM). The work focuses solely on the development and implementation of an Extract-Transform-Load (ETL) pipeline and the harmonization of data structure, without any analysis or use of individual-level data for research purposes. The data used in this process were de-identified on-premise by each contributing hospital in accordance with local data protection regulations and Ministry of Health privacy policies, prior to transformation into the OMOP CDM format. No identifiable data were accessed or processed by the authors.

